# The seroprevalence and trends of SARS-CoV-2 in Delhi, India: A repeated population-based seroepidemiological study

**DOI:** 10.1101/2020.12.13.20248123

**Authors:** Nandini Sharma, Pragya Sharma, Saurav Basu, Sonal Saxena, Rohit Chawla, Kumar Dushyant, Nutan Mundeja, Z SK Marak, Sanjay Singh, Gautam Kumar Singh, Ruchir Rustagi

**Affiliations:** Maulana Azad Medical College, New Delhi; Directorate General Health Services, Government of National Capital Territory, Delhi; State Surveillance Unit, Public Health Wing-IV, Directorate General Health Services, Government of National Capital Territory, Delhi

**Keywords:** Serosurvey, Repeated cross-sectional study, COVID-19, SARS-CoV-2

## Abstract

**Background:** Three rounds of a repeated cross-sectional serosurvey to estimate the seroprevalence and trends of SARS-CoV-2 were conducted from August-October’ 2020 in the state of Delhi in India in the general population aged ≥5 years.

**Methods:** The selection of participants was through a multi-stage sampling design from all the 11 districts and 280 wards of the city-state, with two-stage allocation proportional to population-size. Household selected was via systematic random sampling, and individual participant selection through the age-order procedure. The blood samples were screened using the IgG ELISA COVID-Kawach kit (August Round), and the ERBALISA COVID-19 IgG (September and October) rounds. The seroprevalence was estimated by applying the sampling weights based on age and sex with further adjustment for the assay-kit characteristics.

**Results:** A total of 4267 (n=15046), 4311 (n=17409), and 3829 (n=15015) positive tests indicative of the presence of IgG antibody to SARS-CoV-2 were observed during the August, September, and October 2020 serosurvey rounds, respectively. The adjusted seroprevalence declined from 28.39% (95% CI 27.65-29.14) (August) to 24.08% (95% CI 23.43-24.74) (September), and 24.71% (95% CI 24.01, 25.42%) (October). The antibody positivity was highest in the ≥50 and female age-group during all rounds of the serosurvey, while the decline was maximum among the younger age-group (5-17 years). On adjusted analysis, participants with lower per capita income, living in slums or overcrowded households, and those with diabetes comorbidity had significantly higher statistical odds of antibody positivity.

**Conclusions:** Despite high IgG seroprevalence, there was evidence for waning of antibody positivity with the progression of the COVID-19 epidemic, implying a potential reduction in population immunity, especially if also associated with the lack of trained T cell immunity.

## INTRODUCTION

The COVID-19 pandemic to date has afflicted over 200 countries and accounts for more than 58.1 million cases and 1.38 million deaths [1]. Evidence from seroprevalence studies of the SARS-CoV-2 globally has shown that facility-based surveillance estimation of confirmed cases and deaths lacks population representativeness since most cases have an asymptomatic-mild clinical spectrum [2-4]. Population-based seroprevalence surveys are investigations to screen for antibodies in the blood using a validated testing method to infer the proportion of the population which has been already exposed to the infection and inform as to the likelihood of protection against disease and reinfection [5]. The persistence of an infectious disease is also dependent on the adequacy of the pool of susceptible individuals capable of acquiring and transmitting the infection [6]. The World Health Organization (WHO) therefore recommends conducting sequential seroprevalence surveys to monitor the trends of infection for planning and mounting effective and adequate public health response [7].

Understanding the change in population antibodies for an infectious viral disease like COVID-19 needs to account for the incidence of new infections along with the waning of antibody titres over time when antibody levels fall below detectable threshold levels in the previously infected and recovered individuals [8, 9]. Furthermore, the persistence of the immune response is likely to be modulated by individual characteristics including age, sex, genetics, co-morbidities, prior vaccination, and environmental factors [10]. Consequently, the knowledge gained from seroepidemiological studies is expected to provide key insights in population-level immunity and aid in the implementation of public health interventions towards containing the spread of the COVID-19 pandemic.

Data from large-scale seroprevalence surveys since the onset of the COVID-19 pandemic in UK, Spain, and Iceland have reported varying antibody positivity with the observed reduction in the seropositivity after 3-4 months since the peak of infection [10-12]. Moreover, the rate of decline of the seropositivity has been ascertained to be greater in those with asymptomatic infection compared to those who were diagnosed with real-time PCR. Serosurvey results have also established significant human-human transmission of COVID-19 through individuals with asymptomatic or subclinical infections [13].

In India, a lower-middle-income country with a 1.35 billion population, located in South Asia, over 9 million cases and 0.13 million deaths have been registered due to COVID-19 [14]. A nationwide seroprevalence study conducted in August 2020 reported SARS-CoV-2 antibodies in one out of every fifteen participants [15]. In Delhi, the Indian capital city-state, with ∼19.8 million population, 26,400 cases per million population, and more than 8,000 deaths have been recorded till date [16]. A countrywide lockdown of all educational institutions, marketplaces, and offices including cessation of all public and private transport except essential services was mandated by the Union government from March 23 to June 3 2020 to halt and interrupt the chain of transmission. However, subsequently, gradual relaxation or unlock coincided with a rapid increase in cases, hospitalization, and deaths, with Delhi reaching the first peak of infection in late June 2020 [17, 18].

We report here the IgG seroprevalence and trends of the SARS-CoV-2 infection and their associated sociodemographic factors in the general population assessed through repeated serosurveys at 4-6 weeks interval during August-October’ 2020 in Delhi, India.

## METHODS

### Study design, participants, and settings

We conducted three rounds of a repeated cross-sectional seroepidemiological study from August to October’ 2020. The participants were enrolled from August 1-7; September 1-7 and October 15-21’ 2020. The inclusion criteria were age ≥5 years and residents of Delhi for at least the past six months, while a participant in the previous round of the serosurvey and those medically contraindicated for venepuncture were excluded.

The state of Delhi is located in Northern India and has 11 administrative districts, 274 wards, and ∼250 urban primary health facilities catering to nearly 50,000 population each, distributed uniformly across the state. The proportion of the population as per the settlement type includes slum designated areas (34%), resettlement colonies (12%), authorized and unauthorized colonies (18%), rural areas (12%), and planned colonies (24%) [19].

The primary outcome of this study was the proportion of participants that have serological evidence of SARS-CoV2 infection.

Sampling strategy (Figure 1): comprised of a multi-stage sampling design with two-stage allocation proportion to population size at the district and ward levels. In the August round, within the wards, the participants were selected from the catchment area of the UPHC or Delhi government dispensary. However, to ensure greater sample representativeness, during the September and October rounds, the selection of participants within each ward was as per the proportion of population corresponding to the specific settlement type within each ward. Consequently, the settlements (slum/village/authorized colony/planned colony) within each ward were enumerated by type and selected from this sampling frame by the simple random sampling method. Subsequently, the households within the settlements were identified by systematic random sampling method. Finally, the participants within the households were selected using the age-order procedure in which all eligible individuals were listed in the ascending order as per their age from which one participant was selected through the lottery method.

**Figure 1.**
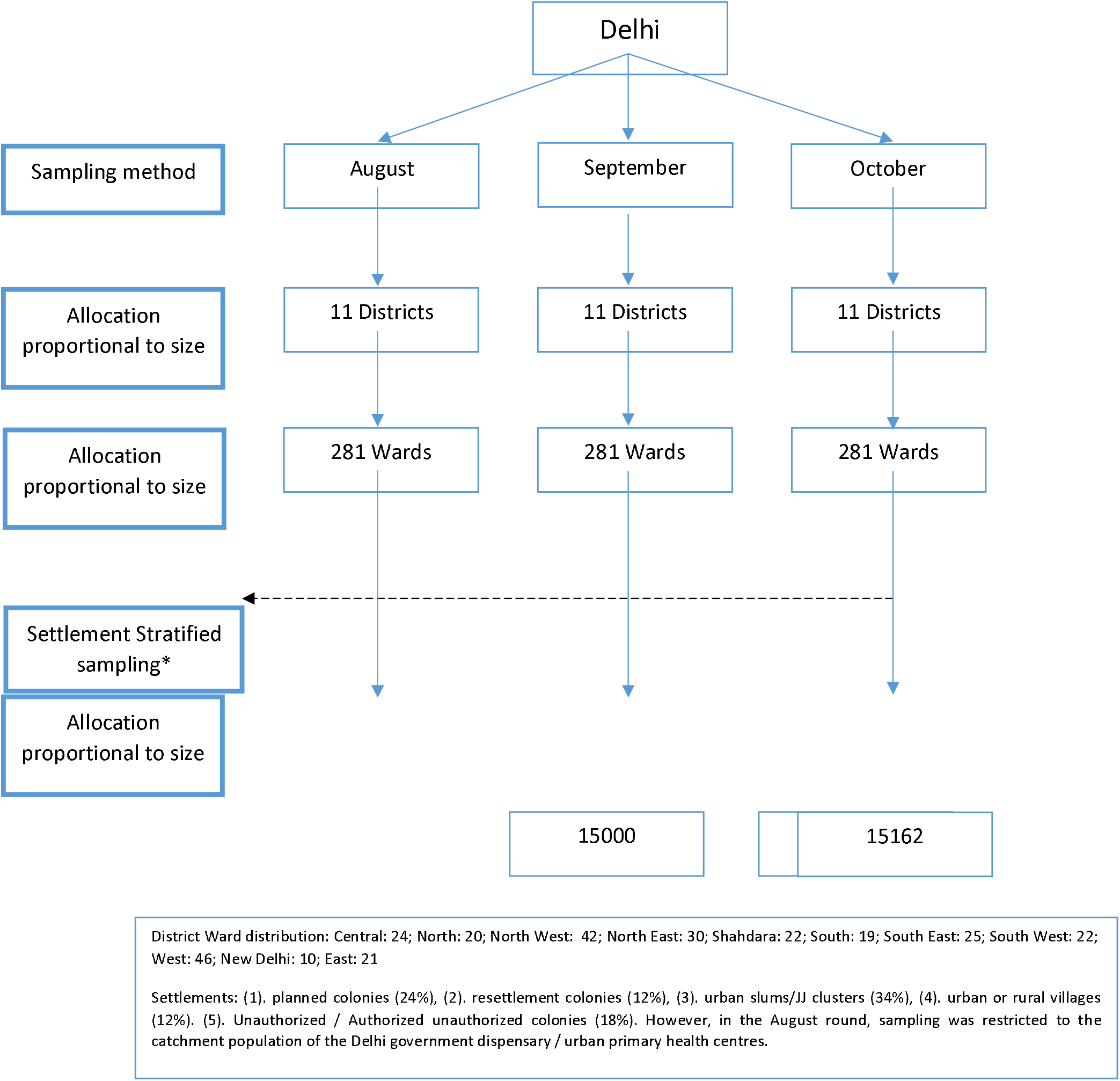
Sampling framework of the Multi stage sampling design.

### Sample size

in the August round was calculated as 15,000 based on the initial July round seroprevalence estimate of 23.48% [20], at 95% confidence levels, 1% absolute precision, a design effect of 2, and accounting for 10% non-response with rounding off. A two-stage allocation proportional to the sample size was applied to estimate the individual district and ward specific sample sizes. From each ward, the participants were selected from the catchment population of the designated UPHC.

### September and October Rounds

The sample sizes for the September and October rounds were based on the district level seroprevalence estimates of the preceding rounds, at 95% confidence levels, and 10% relative precision. The ward sample sizes were estimated from the district sample sizes through allocation proportional to the population size of the respective wards located within their respective districts.

### Procedure

Training and sensitization of survey teams, supervisors, and state nodal officers were conducted a week before the beginning of the surveys. Members of the survey team including frontline and community health workers including the Accredited Social Health Activist, Anganwadi workers, Auxiliary Nurse Midwife, and Public Health Nurse. Around 2- 3 ml, venous blood was collected through venepuncture by a trained phlebotomist or lab technician under all aseptic precautions. The samples were transported to designated government laboratories under standard operating procedures. Data on sociodemographic and clinical characteristics of the participants were collected through face-face interviews conducted by one of the survey team members’ using a pretested, brief interview schedule.

### Kit specifications

The ELISA COVID-Kawach IgG was used in the August round having 92.1% sensitivity and 97.7% specificity [21]. During the September and October rounds, the testing kit used was the ERBALISA COVID-19 IgG with a sensitivity of 99.12% and specificity of 99.33%.

### Statistical Analysis

Data was collected on sociodemographic and epidemiological characteristics of the participants and entered in MS-EXCEL and this data was merged with the serosurvey results data received from the laboratories using a unique identification number that was provided for each participant. The data were analyzed with IBM SPSS Version 25 and Stata 14. The Weighted prevalence was calculated for adjustment of the prevalence by weighing each participant (case) by the inverse probability of their selection to remove the selection bias that occurs from the overrepresentation or underrepresentation of individuals in the sample compared to the population. Age and sex parameters of the population as per census 2011 estimates were applied.

Chi-square was used to assess the significance of the association between the presence of antibodies to SARS-CoV-2 (dependent or outcome variable) and the independent variables (age, sex, overcrowding). The independent variables that showed statistically significant association with the dependent variable were included in a binary logistic regression model. A p-value < 0.05 was considered statistically significant.

The total number of infections was estimated by multiplying adjusted seroprevalence with the estimated population of Delhi. Infection to case ratio (ICR) was calculated by dividing the estimated infection with SARS-CoV-2 confirmed cases by one week and two weeks before the median survey date in each round. Infection fatality ratio (IFR) was calculated by dividing the total number of deaths after three weeks from the median survey date with the estimated number of infections in each round. The estimates of confirmed cases and deaths due to COVID-19 were extracted from the official Delhi state bulletins issued by the government of NCT, Delhi [16].

### Ethical considerations

Written and informed consent for adults and assent for minors was obtained, before recruitment. The study was approved by the Institutional Ethics Committee (F.1/IEC/MAMC/(78/06/2020/No.176).

## RESULTS

A total of 4267 (n=15046), 4311 (n=17409), and 3829 (n=15015) positive tests indicative of the presence of IgG antibody to SARS-CoV-2 were observed during August 2020, September 2020, and October 2020 serosurvey rounds, respectively. There were 439, 202, and 221 samples during the August, September, and October rounds, respectively that could not be processed due to label mismatch, haemolysis, and vial break or leakage.

Antibody prevalence subsequent for weighting for age and sex and after adjustment for the sensitivity and specificity of the test-kits, declined from 28.39% (95% CI 27.65-29.14) (August) to 24.08% (95% CI 23.43-24.74) (September), and 24.71% (95% CI 24.01, 25.42%) (October) (Table 1). Seroprevalence across the districts showed significant variation. The lowest seroprevalence was observed in the South-West district during all rounds of the survey, while the maximum increase in seroprevalence was observed in the Central District (Figure 2A & 2B) (Table 4). The antibody positivity was highest in the elderly (≥50) female age-group during all rounds of the serosurvey, while the decline was maximum among the younger age-group (5-17 years) (Figure 3 & Figure 4).

**Figure 2A.**
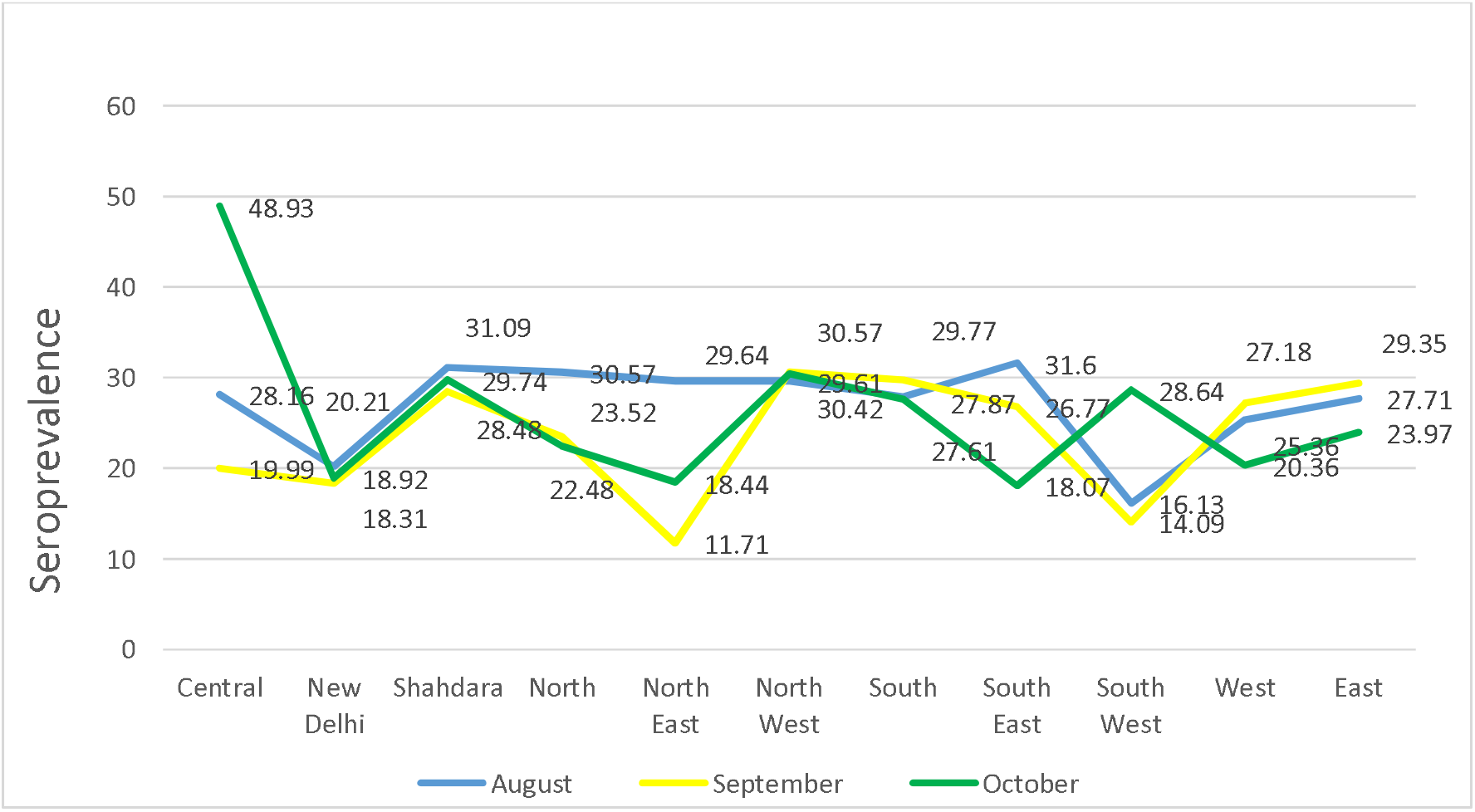
Trends of Seroprevalence of the SARS-CoV-2 IgG antibody in Delhi population from August to October 2020 (NAug;=15046; NSep=17409; NOct = 15015)

**Figure 2B.**
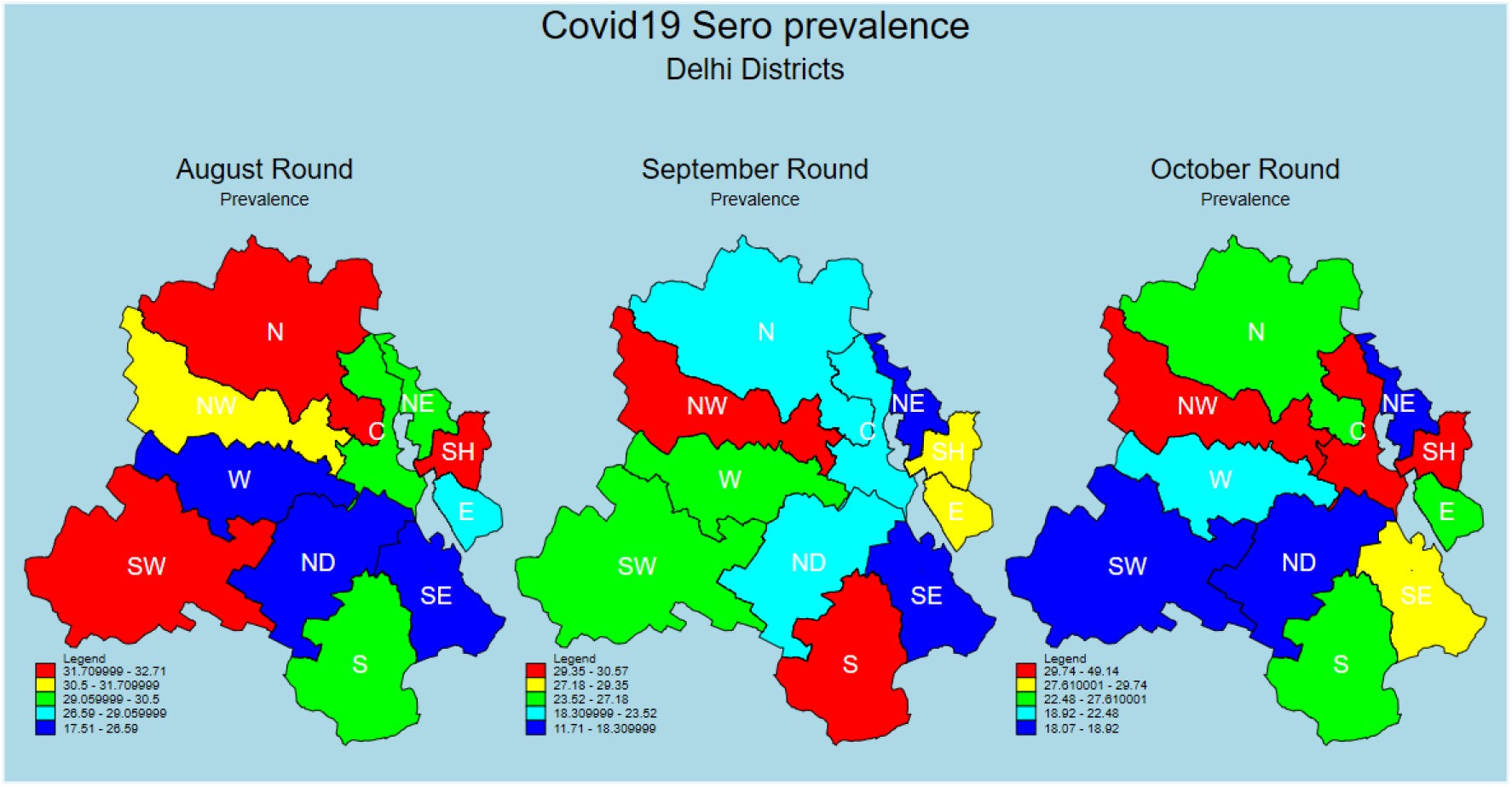

**Table 1.** Comparison of IgG antibody positivity in a repeated cross-sectional study (N=47203)

|  |  |  |  |
| --- | --- | --- | --- |
| 1 | August R1<br>(N=15046)* | Antibody +ve | 4267 (28.35) |
|  |  | Prevalence (Weighted) (95% CI) | 28.84 (28.11, 29.57) |
|  |  | Final adjusted Prevalence (95% CI) | 28.39 (27.65, 29.14) |
| 2 | September R2<br>(N=17409)* | Antibody +ve | 4311 (24.76) |
|  |  | Prevalence (Weighted) (95% CI) | 24.59 (23.95, 25.23) |
|  |  | Final adjusted Prevalence (95% CI) | 24.08 (23.43, 24.74) |
| 3 | October R3<br>(N=15015)* | Antibody +ve | 3829 (25.50) |
|  |  | Prevalence (Weighted) (95% CI) | 25.21 (24.52, 25.91) |
|  |  | Final adjusted Prevalence (95% CI) | 24.71 (24.01, 25.42) |
| 4 | Comparisons | %difference R1-R2 (95% CI) | 4.31 (3.34, 5.28) |
|  |  | %difference R2-R3 (95% CI) | -0.63 (-1.57, 0.31) |
|  |  | %difference R3-R1 (95% CI) | -3.68 (-4.68, -2.68) |

**Table 2.** Socio-demographic characteristics of the study participants (N=46,637)

|  | August R1<br>(N=14779)<br>n (%) | September R2<br>(N=16953)<br>n (%) | October R3<br>(N=14905)<br>n (%) | Total<br>(N=46637)*<br>n (%) |
| --- | --- | --- | --- | --- |
| Age (in Years) |  |  |  |  |
| <18 | 2446 (16.55) | 3013 (17.77) | 2430 (16.30) | 7889 (16.92) |
| 18-49 | 9191 (62.19) | 10544 (62.20) | 9420 (63.20) | 29155 (62.51) |
| ≥50 | 3036 (20.54) | 3392 (20.01) | 3043 (20.42) | 9471 (20.31) |
| Gender |  |  |  |  |
| Male | 7318 (49.52) | 7853 (46.32) | 6955 (46.66) | 22126 (47.44) |
| Female | 7454 (50.44) | 9100 (53.68) | 7948 (53.32) | 24502 (52.54) |
| Education |  |  |  |  |
| Illiterate | 2704 (18.30) | 2012 (11.87) | 1632 (10.95) | 6348 (13.61) |
| Literate | 11784 (79.73) | 14941 (88.13) | 13273 (89.05) | 39998 (85.76) |
| Household size<br>*mean (sd)* | N=14529<br>5.10 (2.42) | N=16534<br>5.17 (10.10) | N=14390<br>4.77 (2.43) | N=45453<br>5.02 (6.91) |
| Income<br>*median (IQR)* | N=12031<br>15000<br>(10000, 25000) | N=13684<br>15000<br>(10000, 25000) | N=13734<br>15000<br>(10000, 25000) | N=39449<br>15000<br>(10000, 25000) |
| Living in urban slum | 2809 (18.46) | 5739 (35.03) | 5981 (39.54) | 14529 (31.09) |
| BPL | 740 (5.01) | 1356 (8.00) | 967 (6.49) | 3063 (6.57) |
| Ever lived in<br>Containment zone | 553 (3.74) | 536 (3.16) | 567 (3.80) | 1656 (3.55) |
| DM | 724 (4.90) | 793 (4.68) | 708 (4.75) | 2225 (4.77) |
| HTN | 709 (4.80) | 671 (3.96) | 659 (4.42) | 2039 (4.37) |
| Urban Slum | 2738 (18.53) | 5872 (34.64) | 5844 (39.21) | 14454 (30.99) |
| Overcrowding | 4051 (27.41) | 4720 (27.84) | 4067 (27.29) | 12838 (27.53) |

\*Missing data due to missing questionnaires, failure to match unique ID and missing data within questionnaires

**Table 3.**
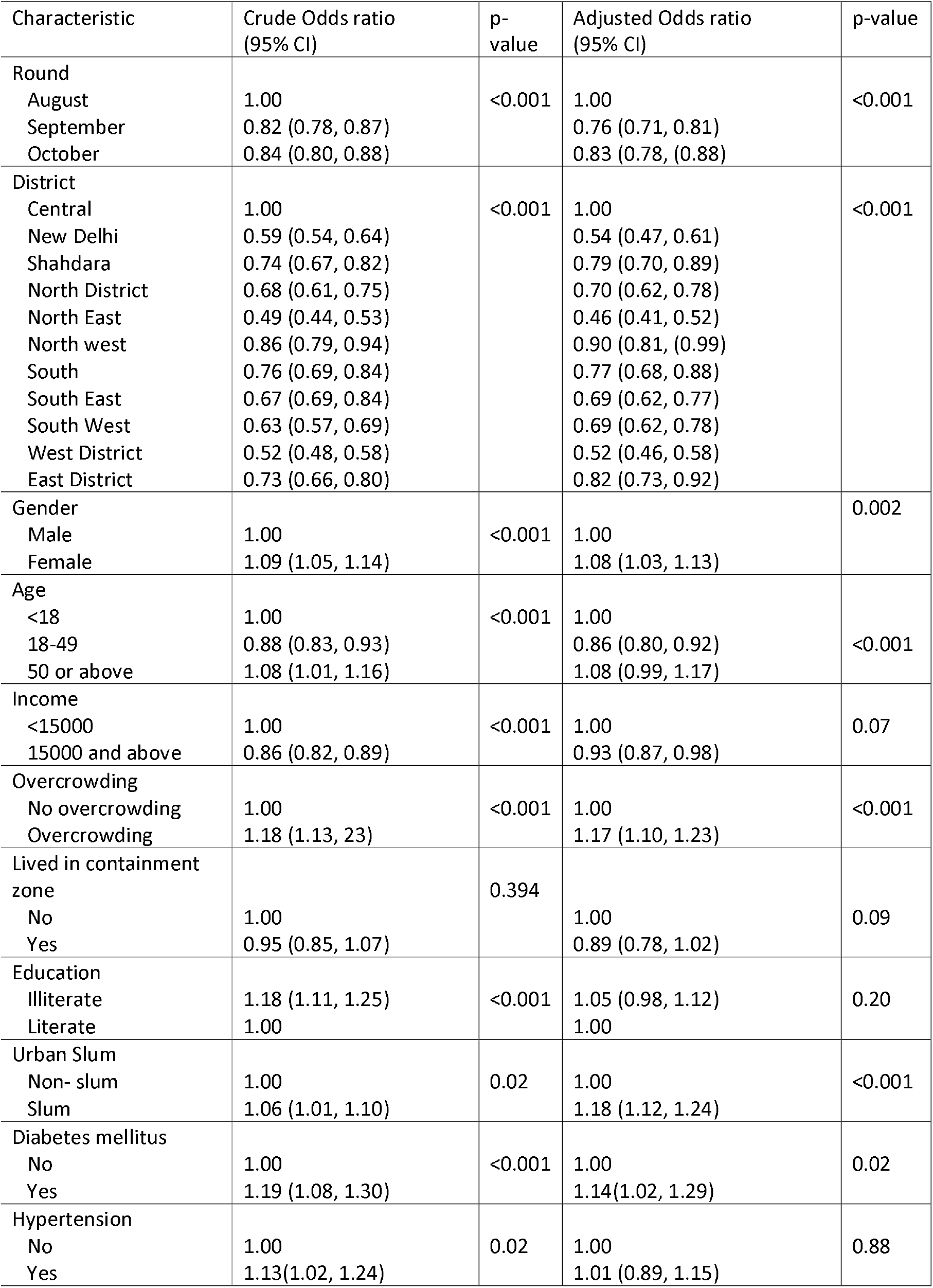
Distribution of factors associated with IgG antibody positivity among participants

| Characteristic | Crude Odds ratio<br>(95% CI) | p-value | Adjusted Odds ratio<br>(95% CI) | p-value |
| --- | --- | --- | --- | --- |
| Round |  |  |  |  |
| August | 1.00 | <0.001 | 1.00 | <0.001 |
| September | 0.82 (0.78, 0.87) |  | 0.76 (0.71, 0.81) |  |
| October | 0.84 (0.80, 0.88) |  | 0.83 (0.78, (0.88) |  |
| District |  |  |  |  |
| Central | 1.00 | <0.001 | 1.00 | <0.001 |
| New Delhi | 0.59 (0.54, 0.64) |  | 0.54 (0.47, 0.61) |  |
| Shahdara | 0.74 (0.67, 0.82) |  | 0.79 (0.70, 0.89) |  |
| North District | 0.68 (0.61, 0.75) |  | 0.70 (0.62, 0.78) |  |
| North East | 0.49 (0.44, 0.53) |  | 0.46 (0.41, 0.52) |  |
| North west | 0.86 (0.79, 0.94) |  | 0.90 (0.81, (0.99) |  |
| South | 0.76 (0.69, 0.84) |  | 0.77 (0.68, 0.88) |  |
| South East | 0.67 (0.69, 0.84) |  | 0.69 (0.62, 0.77) |  |
| South West | 0.63 (0.57, 0.69) |  | 0.69 (0.62, 0.78) |  |
| West District | 0.52 (0.48, 0.58) |  | 0.52 (0.46, 0.58) |  |
| East District | 0.73 (0.66, 0.80) |  | 0.82 (0.73, 0.92) |  |
| Gender |  |  |  |  |
| Male | 1.00 | <0.001 | 1.00 | 0.002 |
| Female | 1.09 (1.05, 1.14) |  | 1.08 (1.03, 1.13) |  |
| Age |  |  |  |  |
| <18 | 1.00 | <0.001 | 1.00 | <0.001 |
| 18-49 | 0.88 (0.83, 0.93) |  | 0.86 (0.80, 0.92) |  |
| 50 or above | 1.08 (1.01, 1.16) |  | 1.08 (0.99, 1.17) |  |
| Income |  |  |  |  |
| <15000 | 1.00 | <0.001 | 1.00 | 0.07 |
| 15000 and above | 0.86 (0.82, 0.89) |  | 0.93 (0.87, 0.98) |  |
| Overcrowding |  |  |  |  |
| No overcrowding | 1.00 | <0.001 | 1.00 | <0.001 |
| Overcrowding | 1.18 (1.13, 23) |  | 1.17 (1.10, 1.23) |  |
| Lived in containment zone |  |  |  |  |
| No | 1.00 | 0.394 | 1.00 | 0.09 |
| Yes | 0.95 (0.85, 1.07) |  | 0.89 (0.78, 1.02) |  |
| Education |  |  |  |  |
| Illiterate | 1.18 (1.11, 1.25) | <0.001 | 1.05 (0.98, 1.12) | 0.20 |
| Literate | 1.00 |  | 1.00 |  |
| Urban Slum |  |  |  |  |
| Non- slum | 1.00 | 0.02 | 1.00 | <0.001 |
| Slum | 1.06 (1.01, 1.10) |  | 1.18 (1.12, 1.24) |  |
| Diabetes mellitus |  |  |  |  |
| No | 1.00 | <0.001 | 1.00 | 0.02 |
| Yes | 1.19 (1.08, 1.30) |  | 1.14(1.02, 1.29) |  |
| Hypertension |  |  |  |  |
| No | 1.00 | 0.02 | 1.00 | 0.88 |
| Yes | 1.13(1.02, 1.24) |  | 1.01 (0.89, 1.15) |  |

**Table 4.** Seroprevalence stratified by districts and serosurvey rounds

| SNO | Districts | August |  | September |  | October |  |
| --- | --- | --- | --- | --- | --- | --- | --- |
|  |  | W | Ad | W | Ad | W | Ad |
| 1 | Central | 29.54<br>(27.18,<br>32.01) | 28.16<br>(25.73,<br>30.69) | 20.36<br>(18.27,<br>22.63) | 19.99<br>(17.89,22.22) | 49.03<br>(46.41,<br>51.66) | 48.93<br>(46.29,<br>51.57) |
| 2 | New Delhi | 21.87<br>(19.11,<br>24.90) | 20.21<br>(17.47,<br>23.17) | 18.72<br>(16.26,<br>21.46) | 18.31 (15.92,<br>20.93) | 19.33<br>(17.39,<br>21.42) | 18.92<br>(17.05,<br>20.92) |
| 3 | Shahdara | 32.42<br>(29.81,<br>35.13) | 31.09<br>(28.47,<br>33.83) | 28.71<br>(26.29,<br>31.26) | 28.48 (26.10,<br>30.96) | 29.98<br>(27.03,<br>33.10) | 29.74<br>(26.91,<br>32.75) |
| 4 | North | 31.89<br>(29.14,<br>34.76) | 30.57<br>(27.82,<br>33.44) | 23.83<br>(21.47,<br>26.37) | 23.52 (21.18,<br>26.02) | 22.78<br>(20.41,<br>25.35) | 22.48<br>(20.14,<br>24.98) |
| 5 | North East | 30.67<br>(27.94,<br>33.55) | 29.64<br>(26.56,<br>32.24) | 12.22<br>(10.48,<br>14.19) | 11.71 (9.98,<br>13.66) | 18.82<br>(17.34,<br>20.39) | 18.44<br>(16.97,<br>19.99) |
| 6 | North West | 30.98<br>(28.96,<br>33.08) | 29.61<br>(27.52,<br>31.77) | 30.78<br>(28.95,<br>32.66) | 30.57 (28.73,<br>32.47) | 30.64<br>(27.52,<br>33.94) | 30.42<br>(27.32,<br>33.70) |
| 7 | South | 29.31<br>(26.65,<br>32.12) | 27.87<br>(25.27,<br>30.58) | 29.96<br>(27.31,<br>32.75) | 29.77 (27.22,<br>32.42) | 27.82<br>(24.76,<br>31.09) | 27.61<br>(24.70,<br>30.67) |
| 8 | South East | 32.87<br>(30.41,<br>35.44) | 31.60<br>(29.17,<br>34.12) | 27.01<br>(24.95,<br>29.17) | 26.77 (24.78,<br>28.87) | 18.46<br>(16.79,<br>20.25) | 18.07<br>(16.42,<br>19.84) |
| 9 | South West | 17.91<br>(15.64,<br>20.44) | 16.13<br>(13.89,<br>18.58) | 14.51<br>(12.83,<br>16.37) | 14.09 (12.44,<br>15.87) | 28.85<br>(26.18,<br>31.67) | 28.64<br>(26.03,<br>31.37) |
| 10 | West | 26.83<br>(24.86,<br>28.90) | 25.36<br>(23.40,<br>27.42) | 27.41<br>(25.66,<br>29.23) | 27.18 (25.44,<br>28.98) | 20.68<br>(18.32,<br>23.25) | 20.36<br>(17.98,<br>22.93) |
| 11 | East | 29.10<br>(26.37,<br>31.99) | 27.71<br>(24.98,<br>30.59) | 29.56<br>(27.05,<br>32.25) | 29.35 (26.79,<br>32.03) | 24.28<br>(21.55,<br>27.23) | 23.97<br>(21.22,<br>26.92) |

W: Weighted; Ad: Adjusted for assay-kit characteristics

**Figure 3.**
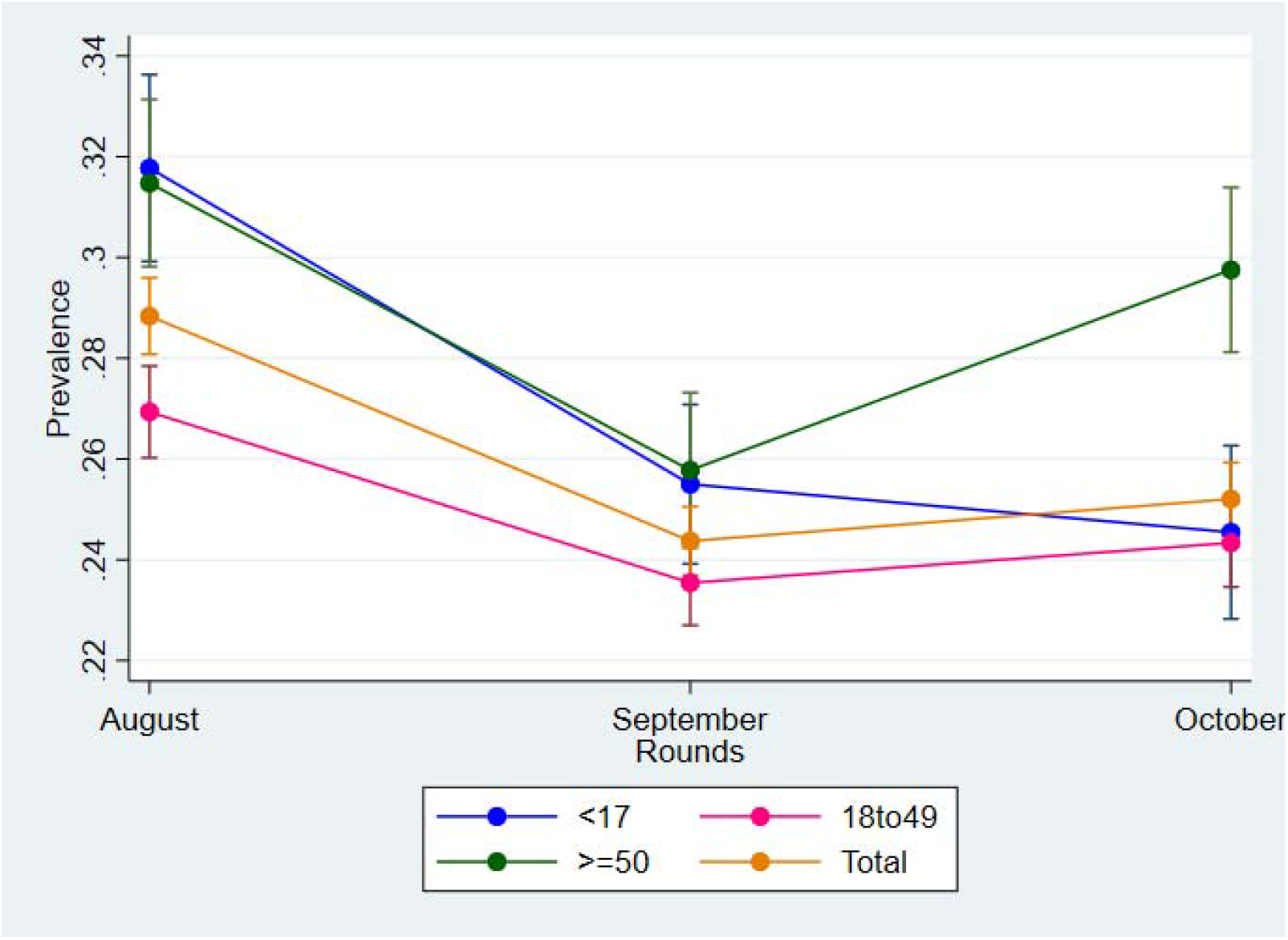
Trends in seroprevalence stratified by age.

**Figure 4.**
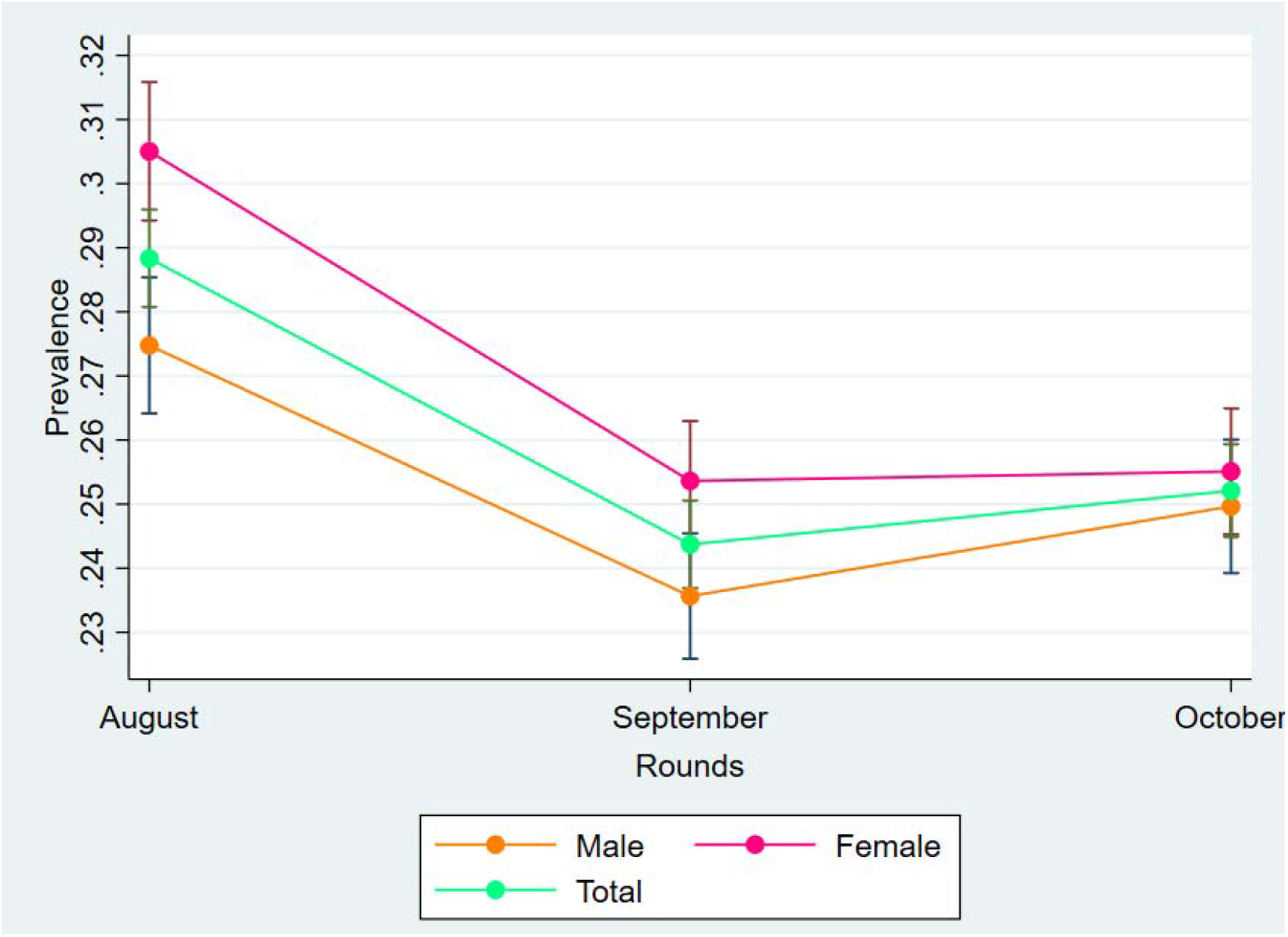
Trends in seroprevalence stratified by gender.

The sociodemographic characteristics of the participants are reported in Table 2. On adjusted analysis, participants with lower per capita income, living in a slum or overcrowded households (three or more people sleeping in the same room), having ever lived in a containment zone and those with diabetes comorbidity had significantly higher statistical odds of antibody positivity (Table 3).

The seroprevalence in those participants with previous COVID-19 infection detected either using the RT-PCR or Rapid Ag Test was 65.2% (n=356), 65.3% (n=226), and 56.5% (n=260), respectively. The point-prevalence of any self-reported influenza-like illness (ILI) among the participants was 4.1%, 9.5%, and 4.4% during the August, September, and October rounds, respectively. The proportion of participants reporting symptoms of ILI and showing antibody positivity was 32.5% (n=575), 27.3% (n=1556), and 25.4% (n=626) during the August, September, and October rounds, respectively.

The infection fatality ratio per 1,000 cases ranged from 0.77 (0.75-0.79) to 0.79 (0.76-0.81), 0.98 (0.95-1.01) to 1.03 (1.00-1.06), and 1.27 (1.24-1.31) to 1.34 (1.31-1.38) during August, September, and October respectively. The infection to case ratio per 1,000 cases ranged from 23.03 (22.91-23.16) to 24.62 (24.49-24.75), 33.05 (32.89-33.21) to 35.98 (35.42-35.75), and 58.95 (58.74-59.16) to 63.33 (63.12-63.55) during the same periods. The population infection projections declined from 5607560 to 4760840 to 4884320 during the August, September and October serosurvey rounds.

## DISCUSSION

The repeated cross-sectional COVID-19 serosurveys in the state of Delhi from August to October 2020 indicated a declining trend in the proportion of participants with detectable antibodies to the SARS-CoV-2 infection. The seroprevalence suggestive of past exposure to SARS-CoV-2 declined from 28.39% (August) to 24.71% (October) although there was considerable inter-district variation in the repeated seroprevalence estimates. These findings concur with the growing recognition of the phenomenon of waning IgG antibody positivity in data from concurrent serial community-based COVID-19 serosurveys globally [9, 10]. Moreover, we observed consistently high rates of antibody positivity in subgroups including those with DM comorbidity, urban slum residents, overcrowded households, female sex, aged ≥50, and amongst previously diagnosed COVID-19 patients. In contrast, antibody positivity declined in the child and adolescent group of participants during subsequent rounds of the serosurvey.

Our findings also corroborate the evidence from early serosurvey reports in Switzerland, the USA, and Canada (April-June’ 2020) which had found the transmission of infection within communities was several times higher as most of the asymptomatic cases of SARS-CoV-2 were not screened using molecular methods [22-24]. Similarly, in this study, 90-95% of participants during all rounds detected with antibody positivity did not manifest any ILI symptoms since 14 days preceding the date of testing, suggestive of extensive asymptomatic seroconversion.

The second round of the national serosurvey in India conducted by the Indian Council of Medical Research during August 2020 reported 7% of India’s population had antibody positivity to the SARS-CoV-2 infection, with an estimated 74 million infections countrywide [15]. The lower seroprevalence observed in the ICMR study was probably because of the considerable variation of the COVID-19 disease burden across Indian states, with Delhi, being one of the worst afflicted states because of its high population density and its large urban-slum population. However, we found the female gender to have a statistically significant positive association with the presence of the IgG SARS-CoV-2 antibody suggestive of differential exposure and susceptibility probably due to behavioural and immunological divergence, respectively. This finding was in contradiction to the nationwide Indian study which did not find any age or sex differentials in antibody positivity [15] but in agreement with the finding of a serosurvey in urban cities of the Eastern State of Odisha in India [25].

Despite the closure of schools and educational institutions throughout the survey, children and adolescents continued to show high antibody positivity implying persistent household level exposure and transmission of infection through mobile adults. This was probable since the period from August onwards coincided with the period of progressive gradual reversal of the nationwide lockdown. Nevertheless, suboptimal adherence to the recommended non-pharmaceutical measures including masking, social distancing, and hand hygiene especially in overcrowded households probably also contributed to the high rates of infection. Our study findings are in agreement with previous serosurvey reports across India which also observed significantly higher odds of antibody positivity in slums and overcrowded households [15, 25-6] which in this study also correlated with the socioeconomic status of the participants.

It is well-established that patients with DM contracting COVID-19 have a clinically poor prognosis and the risk of severe disease manifestation [27-28]. However, we observed the presence of Diabetes Mellitus (DM) comorbidity was also independently associated with the increased risk of subclinical SARS-CoV-2 infection, signifying further accentuation of vulnerability in patients with DM living in slums and overcrowded environments. Consequently, decision making for COVID-19 vaccine prioritization among population subgroups should include both medical (disease) and equity considerations since both factors contribute to the differential risk of infection and prognosis.

In this study, among participants with self-reported recovery from COVID-19 after detection of infection with PCR or Antigen tests, IgG antibodies were detected in only 56.5 to 65.2% cases during the three rounds of the serosurvey. This is even lower than the 81% antibody positivity observed in participants with past SARS-CoV-2 infection during the ICMR serosurvey, highlighting the rapid depletion of IgG antibodies over time [15]. However, European population-based seroepidemiological studies have reported significant variation in the durability of the IgG antibody response post-SARS-CoV-2 infection [10]. Factors such as time since infection, the severity of disease manifestation, and their determinants like age and presence of co-morbidities are likely to influence antibody response. However, the potential for COVID-19 reinfection due to the waning of the antibodies is unclear due to the lack of empirical data from cohort studies with adequate time intervals between the period of infection and prospective assessment. Moreover, the lack of clarity on the role of pre-existing trained immunity acquired and specific T-cell immunity and associated memory responses preclude a clear understanding of the extent of risk of COVID-19 reinfection in those with asymptomatic seroconversion followed by the rapid, progressive depletion of IgG antibodies.

The overall seroprevalence in this study ranging from 24.08%-28.39% with a caveat for the likely underestimation of antibody positivity in subsequent rounds due to the waning of the antibody response in segments of the population, who may or may not have acquired immunity for protection against the infection. Consequently, future studies should estimate the incidence of COVID-19 reinfection in cases with asymptomatic or mild seroconversion which constitutes the clinical profile of most cases to validate the durability of the immune response. The temporal feasibility of such an approach is evident since ≥6 months have elapsed since the initial rounds of population serosurveys were conducted across multiple Indian cities and states. Understanding the phenomenon would help in planning early COVID-19 vaccine deployment through evidence based identification of subgroups for early vaccine coverage prioritization and achieving the minimum herd immunity threshold in the least possible time.

There exist some study limitations. Sample representativeness in the August round was probably lesser compared to the September and October rounds. The sample was underpowered for detecting ward level variations, so disaggregated ward-level data were not analysed. The missing sociodemographic data were assumed to be missing at random type for estimation of the sample statistics. The testing kit did not detect IgM antibodies likely to be present in participants with acute infection but lacking detectable IgG causing underestimation of the population seroprevalence levels.

## Data availability

The anonymized dataset would be made available on reasonable request to the corresponding author.

## Data Availability

The anonymized dataset would be made available on reasonable request to the corresponding author after publication of the article.

